# Sex differences in 1,114,625 varicose vein surgeries: an 18-year cross-sectional analysis of the Brazilian public health system

**DOI:** 10.1101/2025.10.07.25337509

**Authors:** Clara Sanches Bueno, Bruna Castelucci Bongiovanni, Nickolas Stabellini, Stephanye Santos de Paula, Júlia Freire Castanheira de Paiva Ferreira, Bruno Jeronimo Ponte, Felipe Soares Oliveira Portela, Marcelo Fiorelli Alexandrino da Silva, Marcelo Passos Teivelis, Nelson Wolosker

## Abstract

**Background:** Chronic venous disease (CVD) is a highly prevalent condition with significant morbidity. Although there is a well-documented predominance of CVD among women, the differences in surgical treatment based on sex, especially in low- and middle-income countries, are not well understood. This study aims to investigate these disparities within Brazil’s public healthcare system.

**Methods:** This nationwide cross-sectional study analyzed all varicose vein surgeries (n = 1,114,625) performed in the public sector from 2007 to 2024. Data were extracted from the national hospital information system (SIH/DATASUS). We compared demographics, procedural rates, age at surgery, length of hospital stay, and patterns of intercity and interstate travel for surgery between the sexes.

**Results:** Women accounted for the majority of procedures, making up 80.5% of the total. Men underwent bilateral surgeries at a significantly older age than women, with a median age of 49 years for men compared to 47 years for women (p<0.01). Distinct travel patterns based on gender were observed: men were more likely to travel between cities for bilateral procedures (36.84%) compared to women (35.27%, p<0.01). On the other hand, women traveled more frequently between states (0.40% vs. 0.31%, p<0.01). The Southeastern region, which is home to 41.8% of the population, performed the highest volume of surgeries.

**Conclusion:** This study reveals significant gender disparities in the surgical treatment of varicose veins in Brazil, extending beyond mere prevalence to include differences in the age at which intervention occur and geographic access patterns. The findings suggest that biological factors alone cannot account for these disparities, highlighting that healthcare-seeking behaviors, referral patterns, mortality rates, and structural inequities also play a crucial role. Further qualitative research is needed to elucidate the underlying motivations for these differences and to guide the development of more equitable healthcare policies.

## Introduction

Chronic venous disease (CVD) is characterized by dilated, elongated, and tortuous superficial veins in the lower limbs, commonly known as varicose veins. This condition originates from structural deterioration of the venous wall and valvular incompetence, leading to blood reflux and venous hypertension. CVD is a highly prevalent condition, estimated to affect approximately half of the population, particularly adults and older individuals. Beyond its cosmetic concerns, CVD represents a significant public health issue, as its classic symptoms, including pain, heaviness, and edema that worsen throughout the day, substantially compromise quality of life. In more advanced stages, CVD can lead to complications such as venous ulcers, resulting in significant morbidity, healthcare costs, and loss of productivity. (1–4)

The management of CVD, particularly in symptomatic cases or those at risk of complications, often requires surgical intervention. Treatment options have progressed from traditional methods like phlebectomy (stripping) and ligation to minimally invasive techniques.(5) These include thermal ablation using laser or radiofrequency, as well as foam sclerotherapy. (6–9) However, access to these technologies is often inequitable. Within the Brazilian Public Health System (PUBLIC), resource constraints limit the availability of these minimally invasive options, making phlebectomy the standard procedure.

The global prevalence of varicose veins varies significantly, with studies reporting rates ranging from 1% to 75% in the general population. This disparity is due to differences in methodology and population demographics; the condition is reported more frequently in developed countries. For instance, in the United States, varicose veins affect 74.9% of the population. (10)

In Brazil, a low-middle-income country, the PUBLIC plays a crucial role in providing surgical access to 75% of the population. The landscape of CVD in this national context is characterized by high treatment demand, as well as the logistical and resource challenges typical of developing nations. A study analyzing procedures performed within the PUBLIC sector from 2008 to 2019 revealed an average surgical rate for varicose veins of 4.5 per 10,000 inhabitants per year.(11) While this is a relatively high rate, it is still lower than that found in countries like the United States.

International epidemiological studies consistently indicate a significant gender disparity in the prevalence of varicose veins, with the condition being significantly more common in women than in men. Estimates suggest that the prevalence ranges from less than 1% to 73% and from 2% to 56% for males.(12) Studies indicate that these differences may be linked to the influence of hormonal factors, pregnancies, and potential differences in connective tissue composition, which contribute to the underlying pathophysiology. (13–15) However, few studies are exploring whether these differences extend to treatment outcomes.

Despite established knowledge on sex differences in the prevalence of varicose veins, studies are scarce, particularly in low- and middle-income countries, investigating whether this disparity is replicated in surgical treatment. Therefore, this study aims to address this literature gap by investigating whether sex-based differences exist in treatment and describe the epidemiological sex-differences between patients undergoing traditional surgical treatment for varicose veins within the PUBLIC.

### Objectives

This study aimed to conduct a comprehensive assessment of sex-based disparities in 1,114,625 varicose vein surgeries among patients treated within the Brazilian public healthcare system over an 18-year period.

## Materials and Methods

### Data Source

All patient data were obtained from the SIH DATASUS data repository, a platform maintained by the Brazilian Ministry of Health that gathers information about hospitalizations funded by the PUBLIC sector. (16) The raw data underwent an extract, transform and load process, which was created and described in the Fiocruz Applied Health Data Science platform (PCDaS), During this process, data from the SIH DATASUS regarding all hospital admissions nationwide were extracted from the platform and enriched with the integration of other databases. This resulted in a final dataset organized into multiple .csv files by state and year. Subsequently, a query was performed using Python to extract variables of interest from each table and generate a final dataset combining multiple sources. (17) The initial dataset included all patients admitted from 2007 to 2024, although data for the last two months of 2024 were unavailable in DATASUS, specifically for both bilateral and unilateral varicose vein surgeries.

The selection of cases was performed using the procedure codes from the Unified Health System Table of Procedures, Medications, Orthoses, Prostheses, and Materials Management System (SIGTAP/SUS).

The study was approved by the Institutional Review Board (IRB). Since the data are unidentified on the platform, informed consent was not feasible and, as a result, was not requested by the local Ethics Committee.

This study compared various analyses between sexes, including the total number of procedures performed, patient demographics, the proportion of procedures conducted on men versus women, differences in the age at which each procedure was performed, and the length of hospital stay per procedure. Additionally, the study assessed differences in the need to travel between cities and states to undergo the procedure based on sex.

### Covariates

Demographics, admission, and procedure data were obtained for all eligible patients. The demographic characteristics included age, sex (male and female), ethnicity (White, Black, Yellow, Mixed, Indigenous, or others), region of residence within the country (North, Northeast, Midwest, Southeast, and South), as well as the municipality and state of residence.

Admission information included admission type (elective, urgent or other), the municipality and region of hospitalization, the date of admission, the date and status of discharge, and the primary diagnosis ICD code (International Classification of Diseases). Procedure information was composed of the procedure code. (18)

### Data processing and statistical analysis

Data processing and statistical analysis were performed using R (version 4.5.0) with the Cursor integrated development environment (IDE), an AI-assisted coding platform.. (19,20) This tool was employed to create and optimize statistical analysis scripts, facilitating efficient code development for large-scale data processing. Individual state files were consolidated into a unified national dataset. The data cleaning process included age filtering (18-100 years), variable type standardization, and the creation of key derived variables, particularly “travel_status”, which compares the city/states/region of residence to that of hospitalization).

### Variables and descriptive statistics

The study examined demographic variables (sex, age, and ethnicity), clinical variables (admission type and complexity), and healthcare utilization variables (length of stay). We created two outcome metrics: 1) travel status between states and cities, and 2) discharge type (derived from mortality/billing codes).

Descriptive statistics included both absolute and relative frequencies for categorical variables, as well as the median value with interquartile ranges for continuous measures. Since the cost data obtained represent standardized reimbursement rates, no sex-based differences were observed for the analyzed procedures. Consequently, this variable was excluded from our evaluation. To analyze the age difference between sexes, the Bonferroni correction was used to control for type I error in the presence of multiple comparisons.

Data on sex were extracted from the DATASUS database, which primarily sources information from hospital admission records (SIH - Sistema de Informac[ões Hospitalares). Information on sex and ethnicity was obtained through self-declaration at the time of hospital admission. This study is in accordance with the SAGER guideline checklist and STROBE statement. (21,22)

## Results

Between 2007 and 2024, a total of 1,114,625 procedures were performed. The majority (80.5%) were performed on women. The average age was similar for both groups, occurring most frequently in the 5th decade of life. Most patients were identified as White, and the largest proportion of procedures was performed in the Southeast region. The average hospital stay was 2 days for both genders. Bilateral surgeries were more frequent in women, while unilateral surgeries were more common in men. (**Table 1**).

**Table 1.**
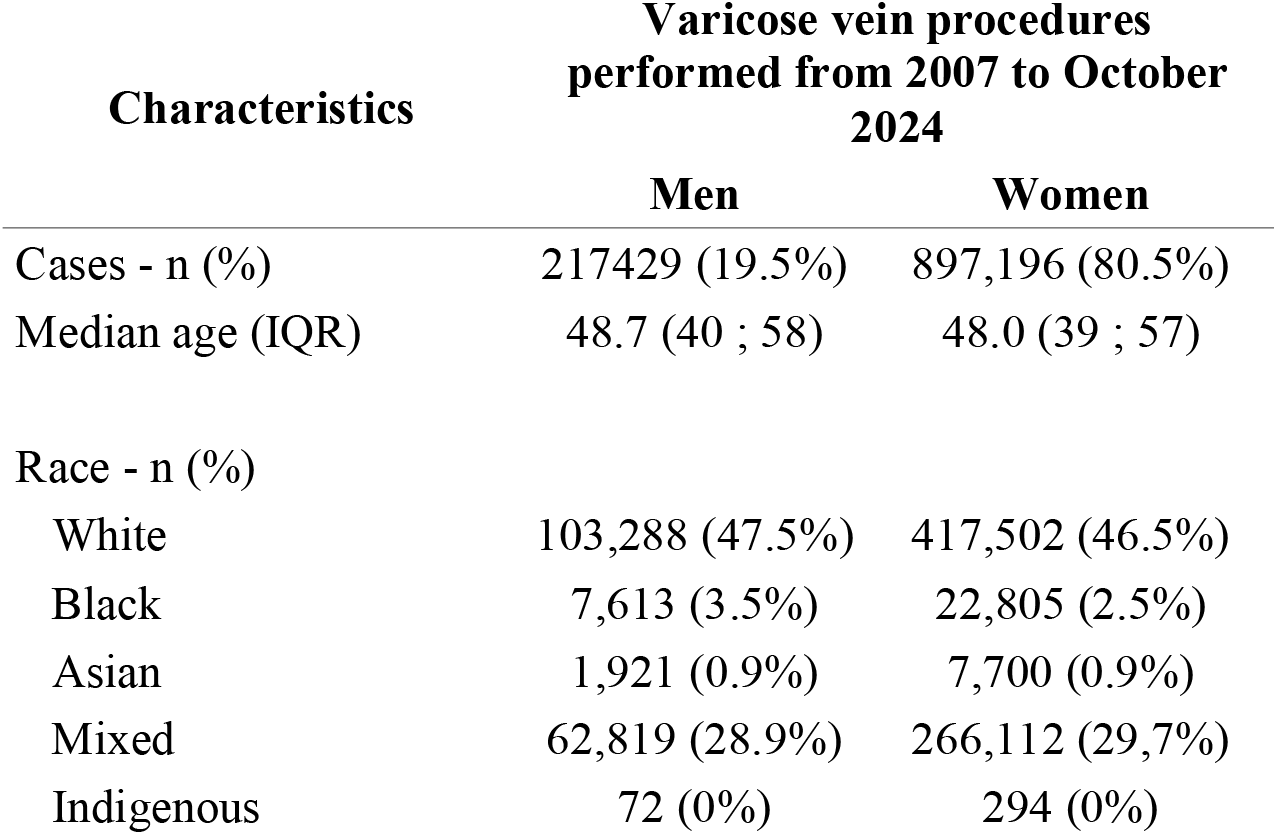
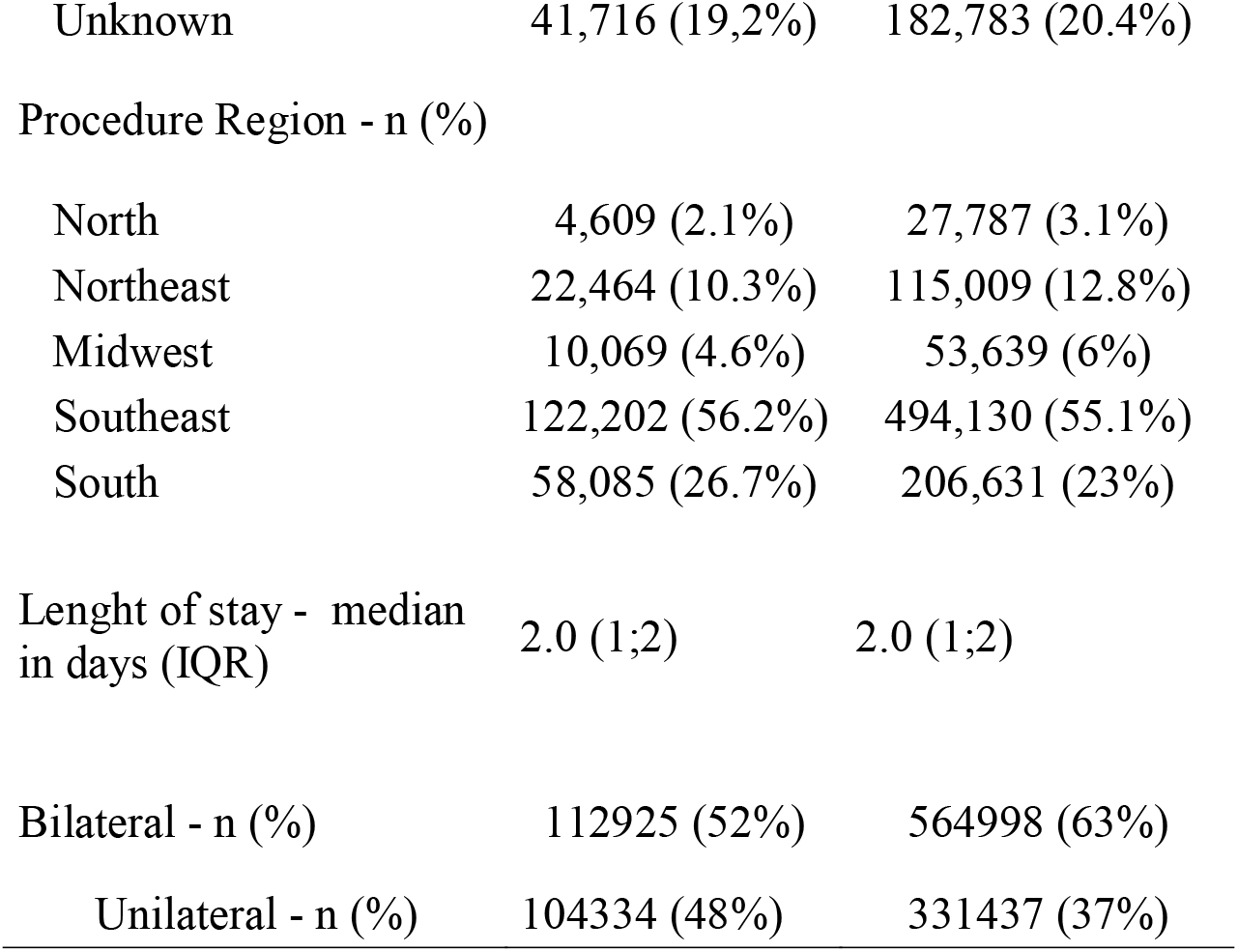
Patient’s Demographic characteristics by gender.

**Table 2** presents gender distribution and age metrics for bilateral and unilateral procedures. There were 678,469 bilateral and 436,156 unilateral surgeries. Women accounted for 83.30% of bilateral and 76.10% of unilateral surgeries. Men underwent varicose vein procedures at a slightly older age than women for the treatment of bilateral varicose veins; for unilateral varicose surgery, there was no statistically significant difference. Also, women underwent most of the procedures.

**Table 2.** Gender distribution and age by procedure.

| Metrics | Bilateral | Unilateral |
| --- | --- | --- |
| Total Surgeries | 678.469 | 436.156 |
| Women surgery - n (%) | 565.454 (83,30%) | 331.742 (76,10%) |
| Men surgery - n | 113.015 | 104.414 |
| Age distribution |  |  |
| Median (Men) | 49 | 49 |
| Standard deviation (Men) | 12,68 | 12,57 |
| Median (Women) | 47 | 49 |
| Standard deviation (Women) | 11,87 | 11.75 |
| Mann-Witney p-value | < 0,01 | 0,344 |
| Bonferroni p-value | < 0,01 | 1 |

Table 3 highlights sex differences in travel for surgery across intercity and interstate distances. A total of 37.9% of patients required intercity travel for varicose vein surgery, amounting to a total of 422,784 journeys. For intercity travel, men travelled proportionally more than women for bilateral varicose vein procedures; however, for unilateral treatment, there was no statistically significant difference. Interstate travel rates were lower, accounting for only 4,243, and Women travelled proportionally more between states than men for varicose vein surgery. (**Table 3**)

**Table 3.** Sex disparities in intercity and in interstate travel for surgery.

|  | Travelers (n) |  | Travel Rate (%) |  | p - value (CI 95%) |
| --- | --- | --- | --- | --- | --- |
|  | Men | Women | Men | Women |  |
| Intercity | 84568 | 333974 | 38.89 | 37.22 | p < 0,01 |
| Interstate | 768 | 3475 | 0.35 | 0.39 | 0.021 |

Mortality rates were low for both sexes, but they were slightly higher for men. There were a total of 64 deaths reported: 43 in men and 21 in women. This corresponds to a mortality rate of 0.0197% for men and 0.0023% for women. Although these rates are low for both genders, men had a rate approximately five times higher than that of women, which is a statistically significant difference (p = 0.0115). The odds ratio for men was approximately 2.02, with a 95% confidence interval.

## Discussion

This cross-sectional study utilizes population data from a publicly available dataset, TabNet (DATASUS), to assess sex differences over 18 years for 1,114,625 varicose vein surgeries. This real-world data study provides insights into epidemiological sex disparities in varicose treatment in a low- and middle-income country with 150.000.000 users of the healthcare system.

The predominance of women in varicose vein surgeries, both unilateral and bilateral, is notable, reaching 80.5% in our study. This may be explained by chronic venous insufficiency and varicose vein disease being significantly more prevalent among women. (23) Hormonal factors, pregnancy, and differences in connective tissue composition are often cited as drivers of this discrepancy. Furthermore, sociological factors likely contribute to this trend, including a greater propensity for women to seek medical consultation for cosmetic or symptomatic concerns related to venous disease. Occupational predispositions, such as prolonged standing in certain professions, may also play a role, as could the use of hormone-based contraceptives and a potential genetic predisposition. (12,24)

White women underwent the highest number of surgical procedures, followed by mixed-race women. Among men, white individuals also had the highest surgical rates. While varicose vein disease is known to be more common among white women, the findings of this study require critical analysis. The disparity in surgical rates should not be interpreted only as evidence of inherent biological differences. It is essential to explore how much of this disparity reflects structural inequities, such as differences in access to specialized healthcare, referral patterns, socioeconomic factors, and the impact of systemic racism, rather than race itself. Further research is needed to quantify the contribution of these societal factors on the observed inequity in surgical care. (7,25)

Multiple studies indicate that men undergoing varicose vein treatment, especially for bilateral disease, tend to be \ older than their female counterpart. This observation is consistent with the results of this study. The difference in age at presentation is largely attributable to distinct healthcare-seeking behaviors between genders. Men often delay seeking medical consultation until their symptoms become severe, influenced by societal norms and cultural perceptions that cause them to overlook early warning signs, like cosmetic changes or mild discomfort. In contrast, women tend to seek treatment earlier for both symptomatic and aesthetic reasons. Consequently, male patients typically present to healthcare providers only after experiencing complications such as venous ulcers, significant pain, or advanced venous insufficiency - conditions that usually develop later in the disease course. (26)

Despite the rarity of death as an outcome from varicose vein surgery, the consistently higher mortality rate observed in men is noteworthy. Although this study did not evaluate contributing factors, it is believed that this disparity may stem from a higher risk profile in men. This profile is often associated with later diagnosis and a more advanced clinical presentation of venous disease, with a higher prevalence of active or healed ulcers, significant edema, and previous thrombotic complications. (27,28)

Additionally, the probable coexistence of cardiovascular and pulmonary comorbidities, such as coronary artery disease and chronic obstructive pulmonary disease, along with a higher rate of smoking, tends to be more prevalent and less managed within the male population. This combination may significantly increase the perioperative risk. (27,29) Therefore, the higher mortality probably is not a reflection of the procedure itself, but rather a convergence of factors related to the severity of underlying diseases and the comorbidity profiles of male patients.

In the Brazilian public healthcare system, the waiting list for elective surgeries, such as varicose vein procedures, can be quite long. Many patients consent to be redirected to other cities or even states within the public network to undergo their procedures more promptly. This necessary travel due to service availability can impact both genders, though the nature of the referrals each gender is willing to accept may differ. Interestingly, men tend to travel more frequently between cities, while women are more likely to travel across state lines. This disparity warrants further investigation to understand the underlying motivations. (30,31)

This gender-based disparity in travel patterns may reflect differences in referral acceptance, access to information, care-seeking behaviors, or the impact of socioeconomic factors. Qualitative research is necessary to investigate the decision-making processes that contribute to these distinct choices.(30,31)

The high volume of varicose vein procedures performed in the Southeast region is not only due to its large population, which accounts for approximately 41,7% of the the country’s total population, but also reflects its crucial role in Brazil’s healthcare system. This can be attributed to the region’s high population density, a greater concentration of specialized professionals, advanced medical services, and its status as the country’s primary hub for technological innovation. (11) Furthermore, the region’s specialized centers attract a significant number of patients from other parts of Brazil who travel there to access higher-complexity care.

## LIMITATIONS

This study has inherent limitations due to the use of the aggregated administrative database TabNet (DATASUS), which is subject to coding errors, reporting inconsistencies, and potential misclassification or underreporting. The data anonymization process also prevents the longitudinal tracking of individual patients, meaning unique patient counts cannot be distinguished from total number of procedures, as one patient may undergo multiple surgeries.

Furthermore, inconsistencies in coding and the quality of reporting make it impossible to categorize patients by comorbidities or clinical severity. This limitation restricts risk-factor analysis and may lead to interpretation challenges and confounding effects in outcome analyses.

Despite these challenges, the strengths of this study are significant. By consolidating Brazil’s public health records, this research provides the most comprehensive real-world analysis of sex-based disparities in varicose vein surgery in a low-to middle-income country to date.

## CONCLUSIONS

This nationwide analysis of 1,114,625 varicose vein surgeries conducted over 18 years reveals sex-based disparities in surgical care across Brazil’s diverse regions.

Women represented the vast majority of procedures, accounting for 80.5% of cases. Mortality rates were low for both sexes, though slightly higher for men (0.0197% compared to 0.0023% for women). Men also underwent bilateral surgeries at a significantly older median age than women (49 vs. 47 years, p<0.01). Notable gender-based travel patterns were observed: men proportionally traveled more between cities for bilateral procedures (36.84% vs. 35.27%, p<0.01), while women more frequently traveled between states (0.40% vs. 0.31%, p<0.01). The Southeastern region, which is home to 41.8% of Brazil’s population, performed the highest volume of surgeries.

## Data Availability

All data produced are available online at DATASUS

https://datasus.saude.gov.br/informacoes-de-saude-tabnet/

## Notes

### Competing Interest Statement

The authors have declared no competing interest.

### Funding Statement

This study did not receive any funding

### Author Declarations

The study used (or will use) ONLY openly available human data that were originally located at:DATASUS (https://datasus.saude.gov.br/informacoes-de-saude-tabnet/)

